# PathFlowAI: A High-Throughput Workflow for Preprocessing, Deep Learning and Interpretation in Digital Pathology

**DOI:** 10.1101/19003897

**Authors:** Joshua J. Levy, Lucas A. Salas, Brock C. Christensen, Aravindhan Sriharan, Louis J. Vaickus

**Affiliations:** Quantitative Biomedical Sciences, Geisel School of Medicine at Dartmouth Lebanon, NH 03756; Department of Epidemiology, Geisel School of Medicine at Dartmouth Lebanon, NH 03756; Department of Pathology, Dartmouth Hitchcock Medical Center Lebanon, NH 03756

**Keywords:** Workflow Automation, Dask, Deep Learning, Whole Slide Images, Segmentation

## Abstract

The diagnosis of disease often requires analysis of a biopsy. Many diagnoses depend not only on the presence of certain features but on their location within the tissue. Recently, a number of deep learning diagnostic aids have been developed to classify digitized biopsy slides. Clinical workflows often involve processing of more than 500 slides per day. But, clinical use of deep learning diagnostic aids would require a preprocessing workflow that is cost-effective, flexible, scalable, rapid, interpretable, and transparent. Here, we present such a workflow, optimized using Dask and mixed precision training via APEX, capable of handling any patch-level or slide level classification and prediction problem. The workflow uses a flexible and fast preprocessing and deep learning analytics pipeline, incorporates model interpretation and has a highly storage-efficient audit trail. We demonstrate the utility of this package on the analysis of a prototypical anatomic pathology specimen, liver biopsies for evaluation of hepatitis from a prospective cohort. The preliminary data indicate that PathFlowAI may become a cost-effective and time-efficient tool for clinical use of Artificial Intelligence (AI) algorithms.

## 1. Introduction

Deep learning is an artificial intelligence method that departs from manual and structural approaches for pattern recognition and prediction by utilizing artificial neural networks (ANN) ^1^. In an ANN, each node receives a weighted signal passed from the output of the previous set of nodes. In each layer, information is aggregated, transformed using a nonlinear function, and passed to the next layer of nodes. Combined with backpropagation (a means of informing weights, connections, filters, etc.), ANNs can be trained to accomplish complex prediction and classification tasks.

Machine learning-augmented digital workflows have the potential to play a significant role in the diagnosis of cancer. A pathologist normally spends hours examining tissue histopathology slides for distinct morphological features indicative of the condition, while unstructured computational approaches aim to assist in this task. The potential future benefits of such workflows in pathology include: decreased time to diagnosis, decreased error rates, reduced burnout, and automation of tedious tasks, among others^2^. Recently, one automated workflow was introduced to classify lung cancer by histology^3^. To achieve widespread clinical use of machine learning tools, optimization of the time and expertise needed to design, train, implement, and interpret such systems will be necessary.

Validation of image analysis technologies by expert review is important in verifying accuracy and in building the trust of the end user clinicians ^4^. Physicians are unlikely to trust a system they cannot comprehend, especially when the results help inform a clinical decision (e.g. aggressive procedures versus palliative regimens). Therefore, transparency and interpretability are key. It is important that clinical machine learning tools are able to “show their work” so end users can better understand the results.

Existing open source workflows such as QuPath have made complex tissue analyses more accessible for the practicing pathologist by the inclusion of user-friendly desktop portals, but these systems, although convenient, may not scale to the high-throughput data demands required for deep learning in the clinical setting^5^. A scalable high-throughput clinical deep learning platform should support applications of different deep learning approaches, without significant manual alterations; training the rich set of models warrants adoption of rapid evaluation methods. For example, mixed precision training^6^, a combination of FP16 and FP32 operations, improves the speed and memory consumption of neural network training (e.g. in a multi-GPU environment (Linux / UNIX), the APEX library from NVIDIA halves GPU memory usage during training with significant performance gains). Usually, deep learning workflows require training image subdirectories housing large swaths of preprocessed images of a fixed size and dimensionality, leading to several downstream problems. First, the directory structure of the image subdirectories is geared towards classification tasks, making the implementation of segmentation algorithms more difficult. Second, a large amount of disk space is needed for these folders and a significant strain is placed on the file management system, especially when transferring folders containing hundreds of thousands of images to other disk locations. Finally, in a clinical scenario, a complete audit-trail of any diagnostic analyses must be retained for quality and risk management purposes. Currently, medical accreditation organizations require every laboratory in the United States to store biopsy slides for a minimum of 10 years^7^; similarly, future medicolegal concerns could require long-term storage of millions of sub-images (or the means to reproduce them).

Evaluation of histology often requires examination of features at multiple different magnifications. The limitations of the traditional image subdirectory paradigm in this context can be appeciated^8^. For example, an investigation of a biopsy from the liver often requires both examination of the inflammatory cells as well as examination of the liver’s microanatomy. The inflammatory cells are best studied at high magnification, because they are significantly smaller than the liver cells. But, evaluation of the liver microanatomy involves differentiating portal (blood vessel, bile ducts, and other connective areas) areas from the liver parenchyma (functional liver cell areas)-tasks best accomplished at low magnification. Existing automated workflows are not amenable to this process. They lack the flexibility to dynamically adjust magnification levels without recreating the set of training sub-images. One solution is to store image tile coordinates (rather than physical sub-images) in a flexible file format. The coordinates could then be accessed using a central SQL repository, in concordance with the Extract Transfer Load framework^9^. Such a system would allow access to any part of the large image at speeds comparable to the less flexible split patch system, while avoiding duplication of the image information.

Liver diseases result in approximately 2 million deaths annually and presents a widespread and growing public health problem^10^. Chronic hepatitides can cause end-stage liver failure. Some affected patients need liver transplantation as a life-saving measure – one that is not always available^11^. Diagnoses of liver disease can be guided by analysis of liver biopsies^12^. The liver is a prototypical example of tissue with two distinct compartments, parenchymal and portal, bearing variable alterations in different disease processes. Different disease processes can be recognized by the spatial distribution of the inflammation, the tissue reaction, as well as the type and degree of damage in these areas. Characterizing these regions is important to assess disease progression and prognosis. A series of scoring systems have been developed to assist pathologists in these efforts ^12–14^. Manual grading and scoring of liver biopsies can be prone to errors of subjectivity, inexperience and bias. Recent computational methods have sought to reduce this burden and automate the classification process, to aid in diagnosis^15–17^.

Here, we present an automated deep learning preprocessing and analytics workflow for classification and segmentation of biopsy morphology, using patch and image-level analysis of whole slide images (WSI). It provides:

1. Rapid preprocessing of slides, utilizing parallelization techniques via Dask^18^.
2. Storage and quick access to slide patches, using a flexible file format that can be read at varying levels of magnification, after storage in SQL
3. Deep learning using mixed precision training^6^ that provides access to many of the most popular classification and segmentation models.
4. Automation of segmentation tasks. This is the first deep learning histology workflow to do this, that we know of.
5. The ability to flexibly perform transfer learning tasks.
6. A visualization framework for interrogating the scoring of the model across the whole-slide image or user-selected patches or Regions of Interest (ROI).
7. Model interpretations using UMAP embeddings ^19^ and SHAP (Shapley feature attribution) ^20^, allowing better clinical insights into the reasons behind the model’s predictions.

## 2. Materials and Methods

### 2.1. Data Acquisition and Processing

Biopsies were obtained via a variety of procedures (percutaneous needle biopsy, transjugular needle biopsy, transgastric needle biopsy, laparoscopic and open wedge biopsy). Samples were fixed in formalin solution, sectioned, oriented and embedded in paraffin. Tissue sections were then cut at a thickness of 5μm and stained with hematoxylin and eosin, using automated staining instruments. Whole slide images at 20X magnification were prepared using a Leica Aperio AT2 scanner. Images were then extracted as SVS files prior to preprocessing.

### 2.2. Description of Framework

The PathFlowAI system provides an end-to-end preprocessing, deep learning, and interpretation command-line framework designed to meet the requirements of a clinical high-throughput digital image analytics pipeline. The seven primary analytic steps are shown in **Figure 1**. We will demonstrate each of these aims on examples of segmentation and classification tasks on liver biopsies for evaluation of steatohepatitis.

**Fig. 1.**
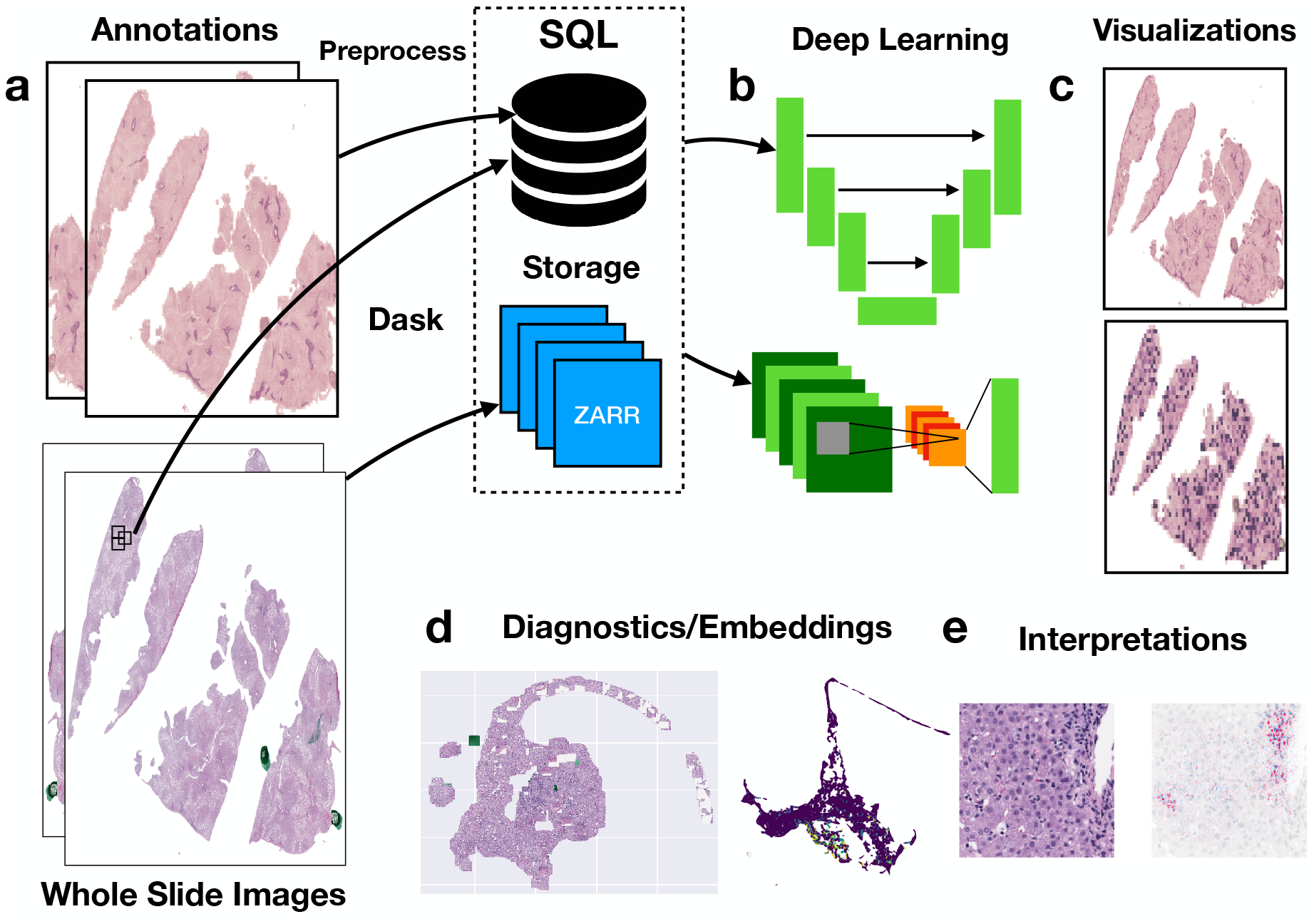
PathFlowAI Framework: a) Annotations and whole slide images are preprocessed in parallel using Dask; b) Deep learning prediction model is trained on the model; c) Results are visualized; d) UMAP embeddings provide diagnostics; e) SHAP framework is used to find important regions for the prediction

#### 2.2.1. Preprocessing Pipeline

The preprocessing pipeline is broken into two sections, accessed through one command in the *preprocessing* module. First, the user supplies whole slide images in common whole slide image formats, including but not limited to SVS, TIFF, TIF, or NPY, and annotation masks for these images in either XML or NPY format. The XML formatted annotations can be produced by a pathologist whole slide image via various annotations software suites (ASAP, QuPath, etc.). The annotations describe polygons encompassing regions of interest of various types (e.g. portal vs parenchyma). The precision of these annotations is not vital, as there is evidence to suggest that deep learning models can learn adequately well from imperfect annotations^21^. The NPY file format is ideal for segmentation tasks, presenting a pixel by pixel classification of each tissue type in an extremely flexible format. The first step of the preprocessing pipeline reads the input whole slide image using Dask and converts the file to ZARR (an array format stored on the disk as a reduced set of compressed, serializable, quick-access chunks), where any specification for an individual patch of the image can be accessed through parallel processes. The masks are retained unaltered if supplied in the NPY format. XML annotations are converted into polygon objects, serialized and stored using python’s pickle format (PKL). At this point, the original image files can be removed to save space. This task is parallelized across all slides through deployment in a high-performance compute cluster.

Next, the pipeline rapidly stores patch level information in a SQL database. The whole slide image is broken into patches of a specified size and marked for inclusion if the intensity of a given patch surpasses a predefined threshold. Patch location, size, and the area of each annotation type in the remaining patches are written to a SQL database. Patch manipulations are done in parallel. Once the information is stored, Dask can be used to rapidly generate image sets using parallel processing. Patches of multiple magnifications are stored efficiently in the SQL database, with one table for each magnification. The storage costs of adding additional magnifications are negligible. The low storage cost allows for experimental data to be retained for long periods of time.

#### 2.2.2. Deep Learning Framework

Once the patch, image and mask data are respectively stored in SQL, ZARR, and PKL/NPY formats, deep learning can be performed on the patch or image level. For segmentation, the NPY masks are loaded using Dask, while patch classifications are supplied from the SQL database. For image-level predictions, the user can optionally add image-level labels as a CSV file.

The *model training* module will train the model on the data using Pytorch, CUDA, and GPUs. The module supports development of classification, regression or segmentation models. The user can select between several classification models (VGG, ResNet, Inception, EfficientNet and AlexNet), and segmentation models (UNET, FCN, Fast-SCNN, and DeepLab). Most of these models are available pretrained, based on features from ImageNet. Transfer learning can also be achieved following pretraining the model on custom datasets. The user can specify many hyperparameters and setup options (e.g. sub/oversampling of particular morphology/annotation) during the training process. It currently does not support an iterative sampling/optimization process. During the training: 1) whole slide images are broken into training, validation, and test sets, 2) each model is exposed to the images from the training and validation sets, with corresponding segmentation masks, and classification/regression targets. Both patch-level and image-level targets are supported. The model performs the prediction and calculates divergence from expectations, referred to as loss between the observed and expected image labels. This loss decreases throughout the iterative training process. When loss of the validation set minimizes, the model parameters are saved. The model with this set of parameters is taken to be the most generalizable model. The validation or test sets of whole slide images can then be scored by specifying a prediction command-line option. This will create an SQL database housing the prediction scores for classification tasks, or segmentation masks for segmentation tasks.

#### 2.2.3. Visualizations

The results of the classification and segmentation tasks are visualized using the *visualization* module. Here, the patches of the whole slide image are colored by the predicted classification. The intensity of each patch is based on the confidence in the prediction. In addition, predicted segmentation masks can be overlaid on the whole slide image and compared to the original segmentation masks overlaid on the same images.

#### 2.2.4. Model Diagnostics and Interpretation

Once predictions have been made, it is useful for the mechanisms of the model to be verifiable and transparent to experts such as scientists, clinicians, and pathologists. This package provides methods for model interpretation using interactive embedding visualizations and SHAP feature attributions. Features that represent distinct categories (which may not necessarily correspond to well-defined histologic categories) identified by the model from the patch level images can be extracted, transformed using UMAP, and then plotted in 3D using the Plotly visualization software. The plots can be labeled using the annotations or classification scores. The degree of cluster separation in these plots can be useful for evaluating how well the model has learned to differentiate the categories involved. In addition, images can be fed into the SHAP framework allowing users to “see” what the model is “seeing” when it makes a prediction. This can be done by overlaying a heatmap on the image. The map reflects how each region in the patch contributes to a particular histologic classification. These SHAP heatmaps can inform classification and segmentation tasks. If unexpected features appear critical to classification, these maps can suggest lines of future inquiry for further investigation of the underlying biology.

### 2.3. Availability

The software is open source, available on PIP as *pathflowai* and on GitHub at https://github.com/jlevy44/PathFlowAI. Contributions are welcome in the form of GitHub issues and pull requests, as the framework is under full development.

### 2.4. Experimental Design

As a proof of concept, liver biopsy for steatohepatitis were evaluated. The evaluation differentiated portal regions from parenchyma and background using both classification and segmentation. Classification tasks were done at the patch level, and segmentation tasks, at the pixel level. Images and masks were preprocessed. An SQL database of data derived from two different levels of magnifications was generated. For the classification task, coarse 512 × 512 pixel patches were used to assess structural differences (portal versus parenchyma), utilizing a ResNet34 ^22^ architecture. For the segmentation task, fine 256 × 256 pixel patches were used to evaluate the exact portal occurrence, utilizing a UNET algorithm^23^. Since the distinction between portal and parenchyma relies on large scale differences, the 224 × 224 pixel patches were not used for that classification. The set of whole slide images consisted of 12 training images, 5 validation images, and 6 testing images. Both the segmentation and classification predictions were overlaid on the whole slide images and compared to the corresponding ground truth labels. The latter were determined by a board-certified anatomic pathologist. The classification patches were embedded and interpreted using SHAP, and the speed of preprocessing was recorded for comparison to other available systems. Since the initial annotations that the pathologist supplied for training were coarse, visualizations of the classification model results were returned to the pathologist to provide high fidelity annotations in regions with higher portal scores.

## 3. Results

### 3.1. Preprocessing Results

The preprocessing pipeline generated patches for 23 fatty liver tissue WSIs at high (224 pixels), medium (256 pixels) and low magnification (512 pixels). In addition, each of the segmentation masks was altered to separate fatty vacuole regions from portal, parenchyma and inflammatory regions (e.g. distinguishing background colored regions within fat vacuoles from true background regions outside of the tissue area). The parallelized framework was deployed on Dartmouth’s Discovery research computing network to process 69 preprocessing jobs (23 WSI x 3 patch magnifications). Patch-level information was stored at no additional storage penalty save for the relatively small generated SQL database. These jobs generated 285,381 patches in 14 minutes. They were divided into high, medium and low resolution, comprised of 144,034, 111,478 and 29,869 patches, respectively. The corresponding results were stored in a 13-MB SQL database and 23 ZARR files (either serialized to form 23 files or stored in 814 uncompressed chunks).

Similar proportions of background/vacuole, parenchyma, portal and inflammatory regions were expected to be found in different WSI, independent of patch size. Across the 23 images, we observed these proportions to be 43.6% background, 49.7% parenchyma, 5.9% portal and 0.9% inflammatory. Importantly, low proportions of inflammatory cells relative to other tissue compartments presents a difficult challenge for image segmentation and classification tasks ^16^.

To illustrate the importance of high-throughput parallelized processing of WSI, PathFlowAI was compared with a slightly modified serial preprocessing workflow ^3^ that utilizes whole slide images. The workflow differs from PathFlowAI in that it breaks up each image into patches and stores them as sub-images in a directory (Fig. 2). PathFlowAI completes the preprocessing task over 16x faster and generates only a SQL database.

**Fig. 2.**
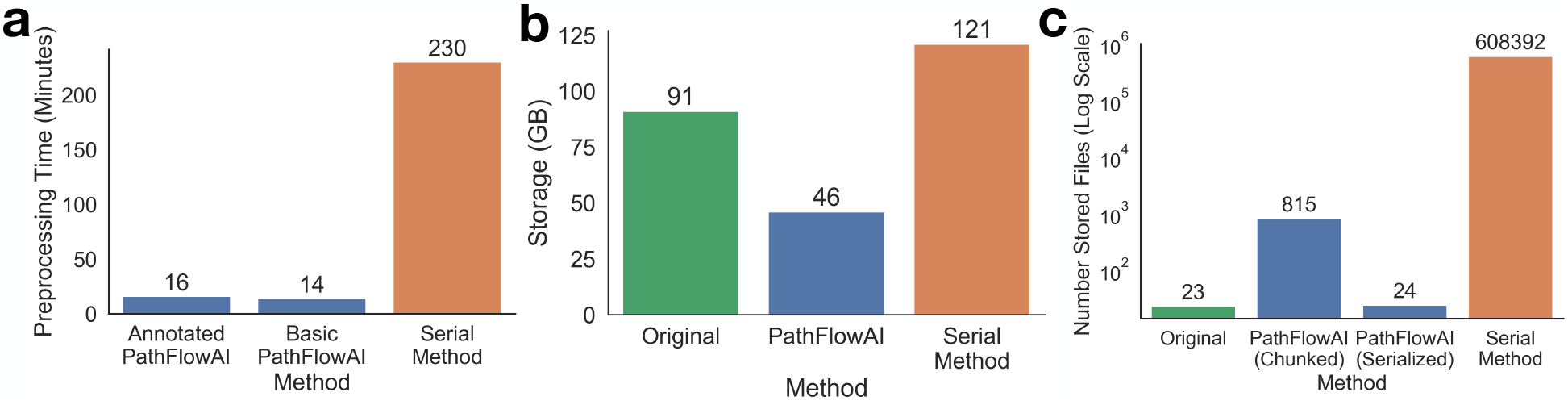
Comparison of PathFlowAI to Preprocessing WSI in Series for: a) Preprocessing time, b) Storage Space, c) Impact on the filesystem. The PathFlowAI method of parallel processing followed by centralized storage saves both time and storage space.

To assess the scalability of PathFlowAI to larger datasets, we independently preprocessed 295 whole slide images of H&E and trichrome stained biopsies. The image set took 52 minutes to preprocess, storage needs were reduced from 1.2 TB to 641 GB, and 10,472 uncompressed ZARR chunks were compressed to 296 serialized files and saved in the file system. Information for 1,024,427 (256 pixel) patches was stored in a 44 MB SQL file.

### 3.2. Segmentation Results

We merged the inflammatory regions with the parenchyma for the medium magnification patches and trained a UNET model to segment portal from parenchyma and background (three separate classes). Overall, the predicted portal segmentations agreed with the physician’s assessment (**Fig. 3a-b)**, with an average sensitivity was 0.71 (**Table 1**).

**Table 1.** Portal Segmentation and Classification Results, Confidence Assessed via 1000-Sample Non-Parametric Bootstrap

| Slide ID | Segmentation |  |  | Classification |
| --- | --- | --- | --- | --- |
|  | Accuracy ± SE | Sensitivity ± SE | Specificity ± SE | AUC ± SE |
| A | 0.93±0.0039 | 0.78±0.028 | 0.93±0.0037 | 0.73±0.014 |
| B | 0.95±0.0031 | 0.79±0.024 | 0.95±0.0029 | 0.77±0.016 |
| C | 0.9±0.003 | 0.67±0.014 | 0.91±0.0031 | 0.85±0.025 |
| D | 0.94±0.002 | 0.77±0.017 | 0.94±0.0019 | 0.78±0.024 |
| E | 0.95±0.0038 | 0.67±0.03 | 0.96±0.0028 | 0.79±0.031 |
| F | 0.92±0.0029 | 0.58±0.019 | 0.93±0.0028 | 0.78±0.011 |

**Fig. 3.**
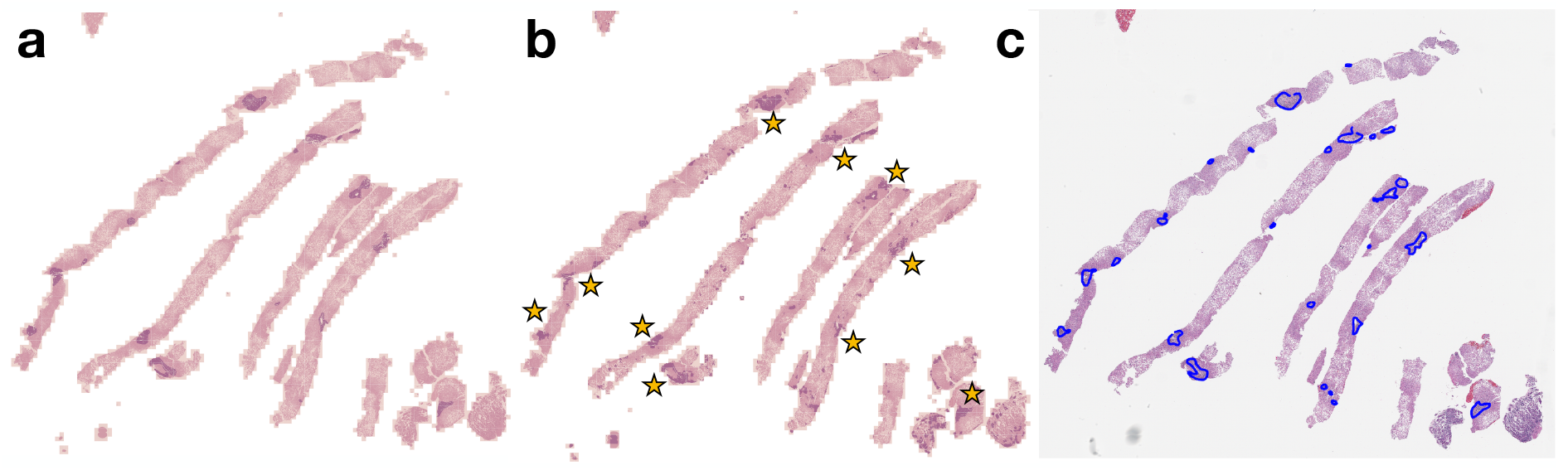
Segmentation: (a) Original Annotations Compared to (b) Predicted Annotations, where Portal, Parenchymal, and Background Regions are Colored Purple, Pink and Orange Respectively; Starred in (b) are Portal Regions that Agree with the Original Annotations; (c) Pathologist Annotations Guided by the Classification Model

### 3.3. Portal Classification Results

Next, PathflowAI was tasked with predicting patches of the image that are more likely to have high concentrations of portal tissue. To train and evaluate the model, a patch was rated as portal if this tissue type covered at least one percent of the patch. The results were presented to the physician and graded. The AUC score was 0.78 (**Table 1**), indicating agreement with physician assessment. The pathologist, blind to the original annotations, was able to more easily annotate the image (**Fig. 3c**) by using results from the patch-level classification model (**Fig. 4a**). Regions assigned a high classification probability tend to overlap with the original pathologist annotations (**Fig. 3a**).

**Fig. 4.**
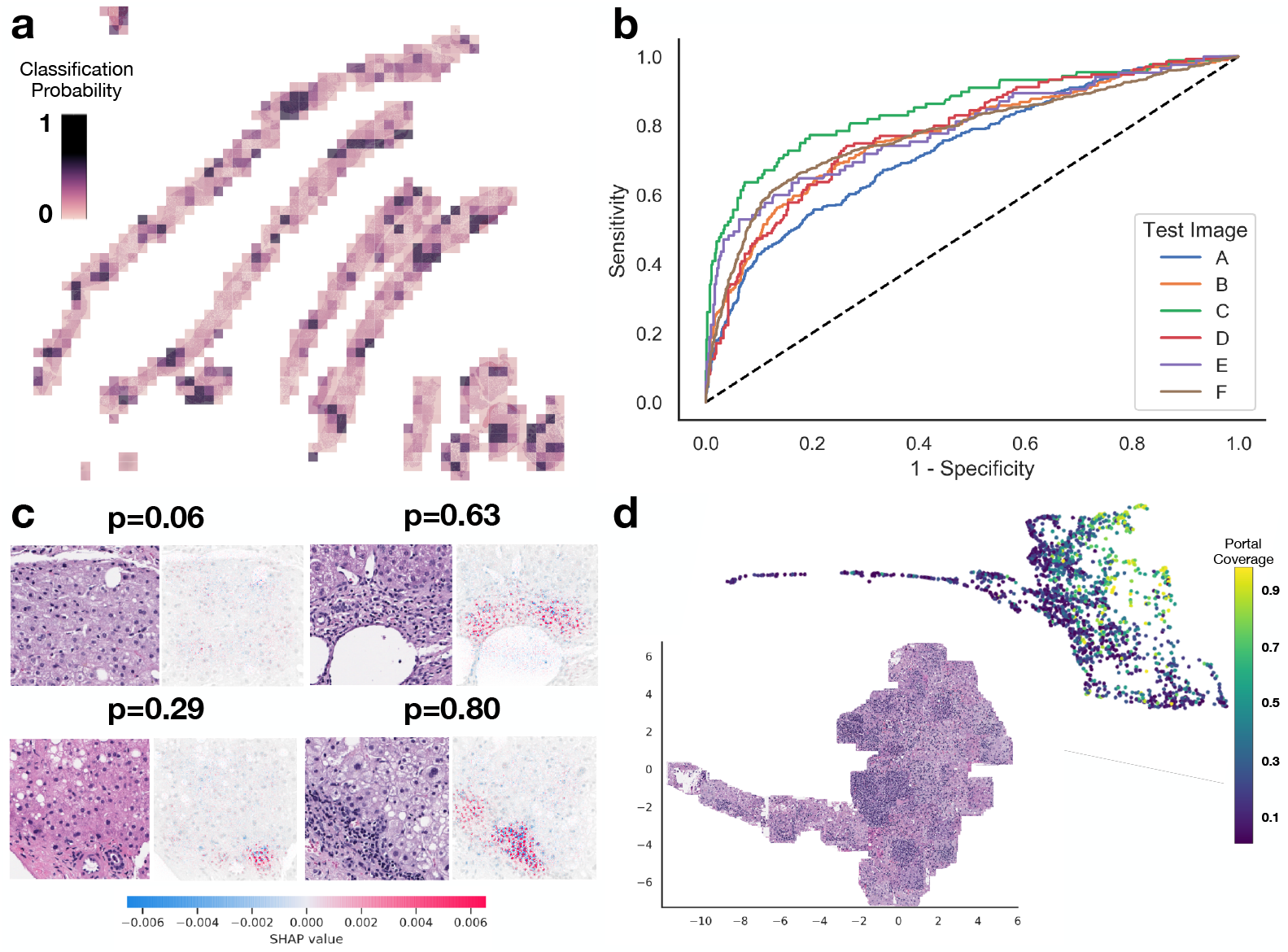
Portal Classification Results: a) Darker tiles indicate a higher assigned probability of portal classification, b) AUC-ROC curves for the test images that estimate overall accuracy given different sensitivity cutoffs, c) H&E patch (left) with corresponding SHAP interpretations (right) for four patches; the probability value of portal classification is shown, and on the SHAP value scale, red indicates regions that the model attributes to portal prediction, d) Model-trained UMAP embeddings of patches colored by original portal coverage (area of patch covered by portal) as judged by pathologist and visualization of individual patches.

### 3.4. Diagnostics and Interpretation

Finally, significant diagnostic information for portal patch predictions were assembled, in the form of embeddings and SHAPley attributions. The embedding plots in **Figure 4c-d** demonstrate class separation and clustering of similar morphology. The SHAP plots reveal regions responsible for the prediction. The algorithm found that features preferentially associated with the periportal category include the presence of lymphocytes. This is congruent with published expertise on the subject^24^. As the prediction score increased for portal classification, SHAP was more likely to assign regions with higher concentrations of inflammatory foci to the periportal category.

## 4. Discussion

We present PathFlowAI, a digital pathology image analysis suite capable of handling the high-throughput workloads typical of clinical practice. Our initial results demonstrate an efficient and flexible pipeline that enables rapid design, training, testing and implementation of deep learning tools. For each WSI, the preprocessing took minutes; this method was at least 14 times faster than the evaluated serial preprocessing method. Additional tests on a larger group of images verified that the required time and storage for preprocessing scale with sample size. In a typical midsized academic laboratory, such as Dartmouth Hitchcock Medical Center, with an estimated volume of approximately 35,000 annual cases, use of PathFlowAI for one year would be expected to reduce storage space required by 38.1TB, the number of files by 308.6 million and preprocessing time by 1,826 hours (76 days, 228 eight-hour workdays) with current resources. Lower resolution images would necessarily be processed even more rapidly.

A proof-of-concept application for basic tissue classification and segmentation tasks on liver biopsies performed as expected. Our preliminary data indicate that PathFlowAI can be effectively used to develop future classification and segmentation algorithms, not only in liver pathology but in diseases involving other organs as well.

Limitations of this study include lower than expected sensitivity and the inclusion of a small training, testing and validation set for the deep learning tests. In addition, although we intend PathFlowAI to be used in clinical scenarios, we have not yet carried out validation trials for its use in this indication. Neither have the models employed by this workflow been optimized for the ideal neural network architecture, hyperparameters, and data augmentation/sampling techniques. The model generated by the pipeline showed reasonable portal region sensitivity in both classification and segmentation tasks but was substandard for a usable clinical diagnostic aid because of the model’s low sensitivity. Some of this could be accounted for by imperfect physician annotations; also conceivable is correct scoring of tissue missed by the physician; both are likely to occur when the pipeline is rolled out into practice. We acknowledge that multiple pathology reviewers will add robustness to this approach. Finally, interpretations using SHAP indicated that the model seemed to consider inflammation as a significant feature of a “periportal” region, which is a point of nonrandom systematic bias in the overall study design. However, “seeing” what the model “sees” presents an excellent opportunity to identify this bias and pre-empt sources of error. Thus, the purpose of this study was to demonstrate several classic procedures that might be employed to create a similar clinical algorithm, not to produce a highly accurate liver tissue processing algorithm.

In the future, the workflow could be applied to other diagnostic tests, such as X-rays, CT images, and MRI scans. Incorporation into standardized, reproducible and scalable frameworks such as Docker ^25^ and Common Workflow Language ^26^ may further make this workflow high-throughput and clinically tractable. We also intend to implement robust QC measures, such as those developed for popular frameworks like HistomicsTK ^27^ and HistoQC^28^.

## 5. Conclusion

Flexible, modular, interpretable, and fast workflow development is necessary for the widespread use of AI in medical practice. Here, we demonstrate PathFlowAI’s ability to preprocess images with minimal storage space and optimal use of time. PathFlowAI is able to rapidly furnish preprocessed data with many different types of ANNs and interpret deep learning methodologies with minimal coding. We hope that this open-source software will facilitate the use of AI in clinical medicine and foster meaningful improvement in the practice of digital pathology.

## Data Availability

Raw data was generated at Dartmouth Hitchcock Medical Center. The group data that support the findings of this study are available upon reasonable request from the corresponding author, JJL. Individual anonymous level data may be available by application to the relevant institutions after obtaining required IRB approvals.

## 6. Acknowledgements

This work was supported by NIH grant R01CA216265. JL is supported through the Burroughs Wellcome Fund Big Data in the Life Sciences at Dartmouth. We would like to acknowledge Christian Haudenschild for help with the code review. *Preprint of an article published in Pacific Symposium on Biocomputing © 2020 World Scientific Publishing Co*., *Singapore, http://psb.stanford.edu*.

